# Family Presence and Audience Effects on Cardiopulmonary Resuscitation: Perceptions of Pre-hospital Emergency Caregivers in Turkey

**DOI:** 10.1101/2020.01.21.20017988

**Authors:** Hasan Erbay

## Abstract

**Background:** There have been many studies regarding family presence during cardiopulmonary resuscitation and most of them are about emergency services or intensive care units. However, the issue has not been studied enough in terms of pre-hospital emergency medicine and the perspective of pre-hospital emergency caregivers.

**Aim:** In this study, it is aimed to present the perceptions and attitudes of a group of pre-hospital emergency care professionals to family presence during cardiopulmonary resuscitation.

**Method:** The data for this descriptive research was collected between March-May 2015. The participants were 63 pre-hospital healthcares from a small city, Afyonkarahisar. The data was collected using a questionnaire and the responses summarized by using frequencies and percentages. Descriptive statistics were used to characterize the sample and each of the survey items.

**Findings:** Of the respondents, for demographic findings of each groups by highest rates; 38 (60.32%) were male, 31 (49.21%) were paramedics and 37 (58.73%) were 1-3 years work experienced. 65.07% were strongly opposed to the family presence during CPR. Among the total sample, 71.41% of participants did not agree with that the presence of press members positively affects their performance.

**Conclusion:** The presence of significant others during CPR affects pre-hospital emergency caregivers negatively. Disturbing effect on caregivers is related not only to the presence of family members or to other significant others but also related to the press and audience. Family presence causes health professionals to experience performance anxiety. Family presence and the audience effect are crucial issues that need more attention in pre-hospital cardiopulmonary resuscitation.

## Introduction

Cardiopulmonary resuscitation (CPR) includes performance of chest compressions, airway management, rescue breathing, rhythm detection, and shock delivery (if indicated) by an integrated team of highly trained rescuers who are competent both for in-hospital and out-of-hospital settings to the casualties thought to be in cardiac arrest[1]. Pre-hospital emergency caregivers (PECs) are confronted with a number of ethical considerations when they are on their way to treat a person who suffers an out-of-hospital cardiac arrest with CPR[2,3]. One of the conflicts in the pre-hospital setting is family-witnessed CPR called family presence during CPR (FPDR)[4–7].

It is a long-running debate and family presence during resuscitation means that the family members can witness the visual and/or physical contact of the caregivers with the patient during resuscitation[8]. There are several aspects of FPDR that concern the patient, their relatives, and emergency care providers[9,10]. In many studies, this issue has been evaluated from the perspective of the family members and it has been shown that they mostly have positive views on FPDR[11–13]. Many specific studies also have investigated the perceptions of health care professionals toward FPDR and they have presented various involvements of it revealing that the issue is not so clear from their side, and it contains many discussions[12,14]. Although, many associations and councils such as like The Emergency Nurses Association (ENA) (2009), European Federation of Critical Nursing Associations, European Society of Cardiology Council on Cardiovascular Nursing and Allied Professions, American Heart Association[15], and European Resuscitation Council (2010) Guidelines for Resuscitation[16]support and recommend in-hospital FWR; for the most part, no guidance exists around how to support family members best in the pre-hospital settings[17].

However, FPDR is a controversial issue because of its ethical, cultural and legal aspects and its effect of psychological or emotional trauma on the patient’s relatives[18–27]. Moreover, discussions about FPDR are almost for in-hospital CPR[28] (in emergency room and intensive care units); not about pre-hospital CPR. Most studies focus on the perceptions of physicians and nurses in hospital, and there is not many things reported about the opinions of other CPR team members including paramedics, emergency nurses and emergency technicians. Unfortunately, the results are quite limited in terms of this issue from the perspective of pre-hospital emergency healthcare workers[29]. PECs are at the forefront of providing immediate CPR to the patient usually in patient’s home, and yet little is known about their experiences and perceptions of FPDR; whereas, family witnessing is sometimes inevitable in pre-hospital practice.

At this point, this study aims to present the perceptions and attitudes of a group of pre-hospital emergency care professionals regarding the family presence during cardiopulmonary resuscitation in Turkey. The term “family” in this paper, not only refers to close family members of the patient, but also the significant others that means friends, neighbors or colleagues at the scene of CPR attempt.

### Design

This was a descriptive study carried out between March–May 2015. Before starting the questionnaire, a preliminary study was conducted on 12 participants and some of the statements in the form were rearranged. Then, the study took place in a small city Afyonkarahisar, Turkey. Eighty-our healthcare providers on active duty on their shift were enrolled in this study. Eight declined to participate, seven forms were filled incomplete, and six forms were excluded because of answers were inconsistent. Overall, final evaluations were made on the forms of sixty-three participants.

### Data collection

Data was collected using a structured questionnaire developed by the authors. It consisted of two parts; the first section collected demographic data (gender, role and year of experience) and the second section was consisted of 18 items each was scored on a 11-Point-Likert-Scale. Each participant was asked to evaluate the content of the questionnaire by evaluating each item on an eleven-point scale: 0 = absolutely disagree, 5 = no opinion/not sure and 10 = absolutely agree.

### Data analysis

The 18 expressions in total from the data collection forms were re-evaluated by considering similar characteristics and rearranged in 10 items. Data was analyzed using the Statistical Package for Social Scientists (SPSS, version 11.0 for windows: SPSS Inc., Chicago, IL, USA). Statistical advice was sought from a statistician to analyze the data. Descriptive statistics were used to characterize the sample and each of the survey items. Mann Whitney U Test was used to compare differences between gender, and the Kruskal-Wallis was used to check the differences between role and year of experience that the samples are independent.

### Ethical considerations

The approval of the study was obtained from the local Ethics Committee. The participants were informed of the purpose of the study, were assured of their right to refuse to participate or to withdraw at any stage and that data would be anonymous.

## Results

Among the 63 responders, for demographic findings of each groups by highest rates; 38 (60.32%) were male, 31 (49.21%) were paramedics and 37 (58.73%) were 1-3 years work experienced. The demographic findings are summarized in Table 1.

**Table 1.**
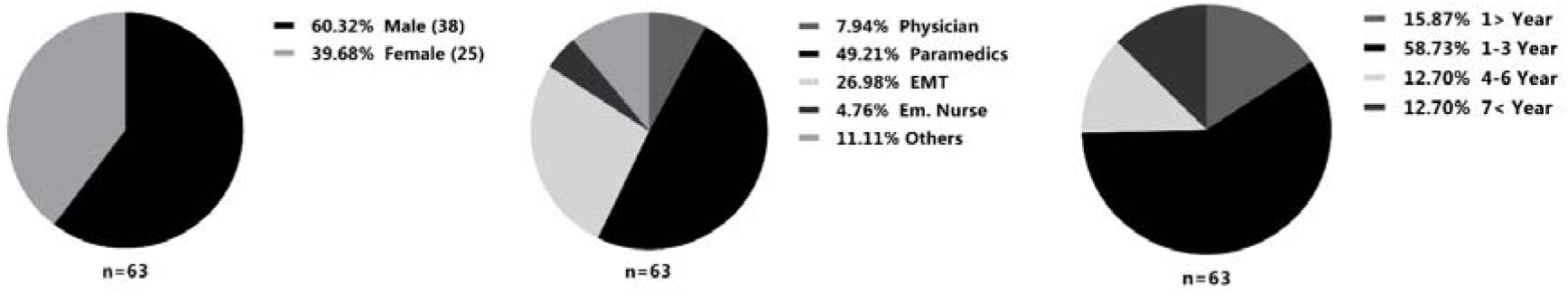
Demographic Characteristics of Participants.

Responses of participants for ten phrases are given in Table 2-11. Among the 63 responders, it is given number of the participants, mean and standard derivation (SD) of the response.

**Table 2.**
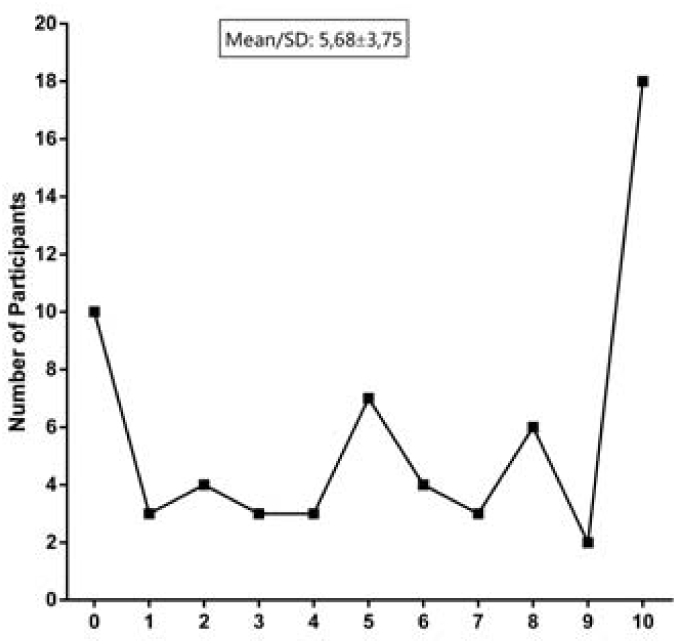
I don’t want to do CPR while people watching me.

**Table 3.**
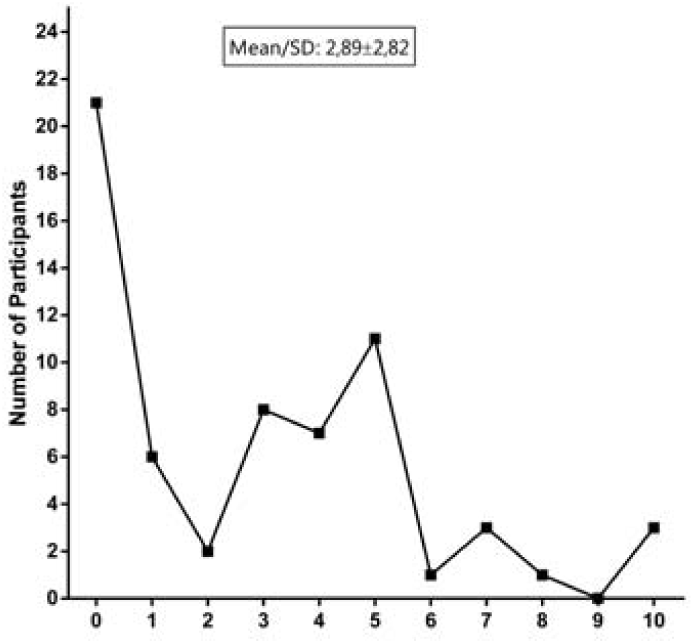
Having someone watching me while applying CPR to a patient, positively affects my performance on CPR.

**Table 4.**
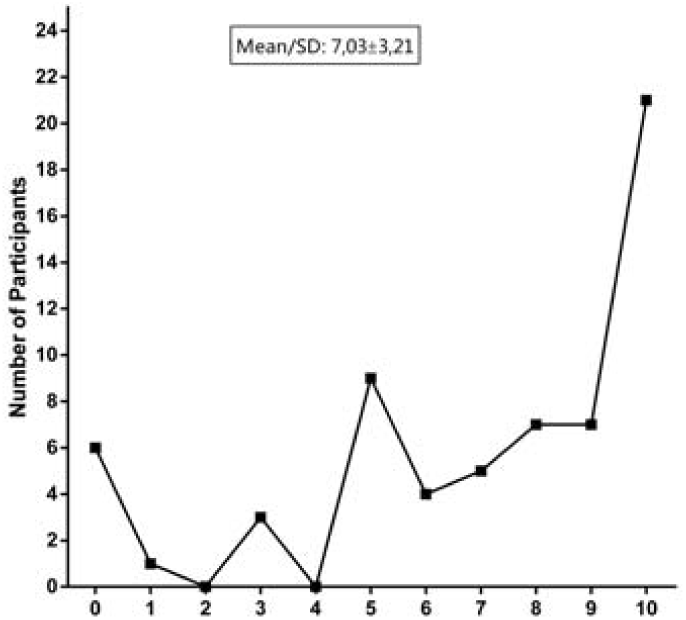
I think, having someone around watching them while an ambulance crew is performing CPR to the patient, negatively affects the performance of the team.

**Table 5.**
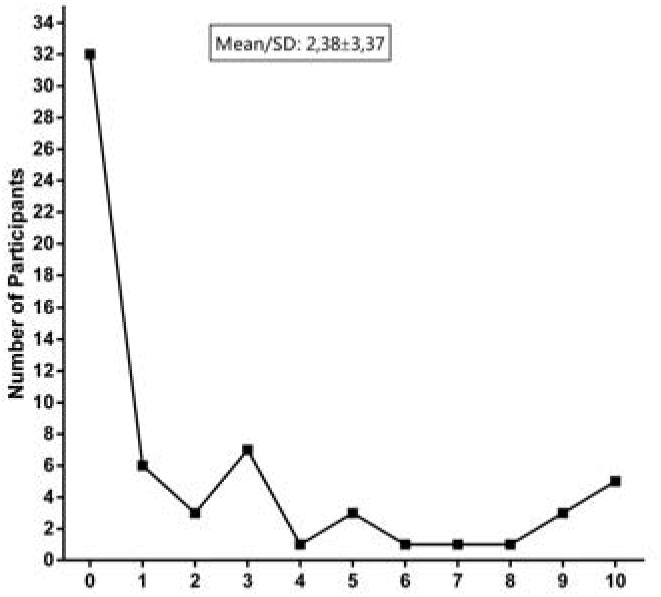
When applying CPR to a patient, relatives can be present and witness our intervention if they request.

**Table 6.**
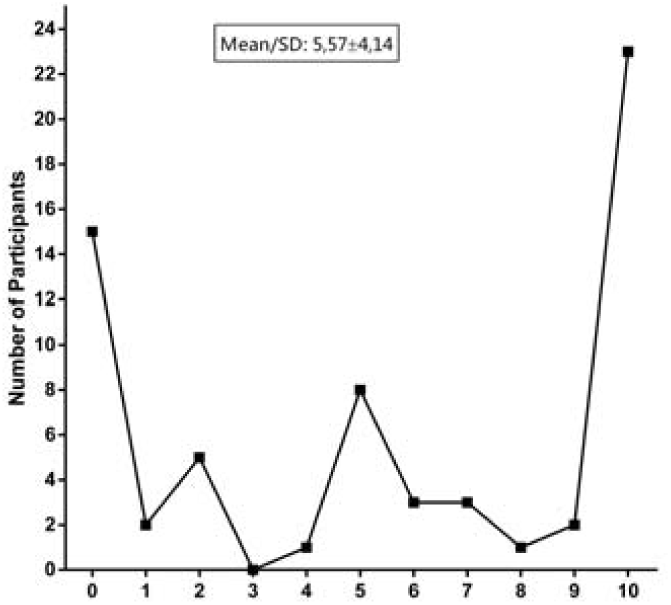
I think I would like to be there if I have the opportunity when being applied CPR to someone close to me.

**Table 7.**
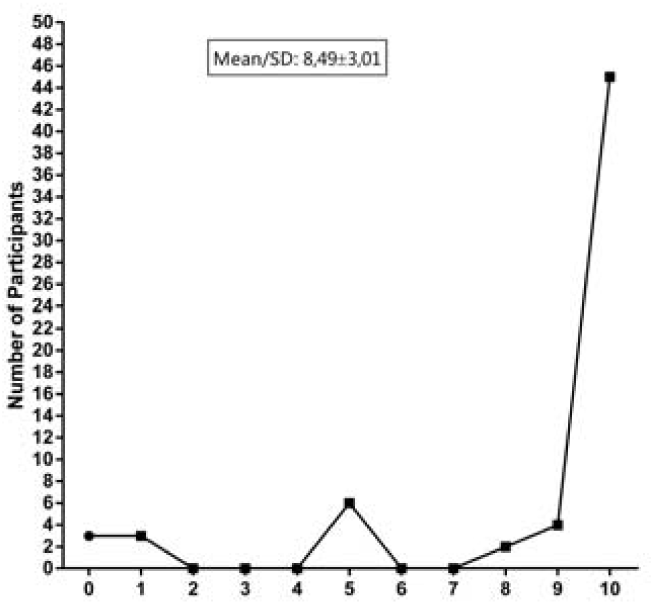
I don’t want any press member to be there when I perform CPR on a patient.

**Table 8.**
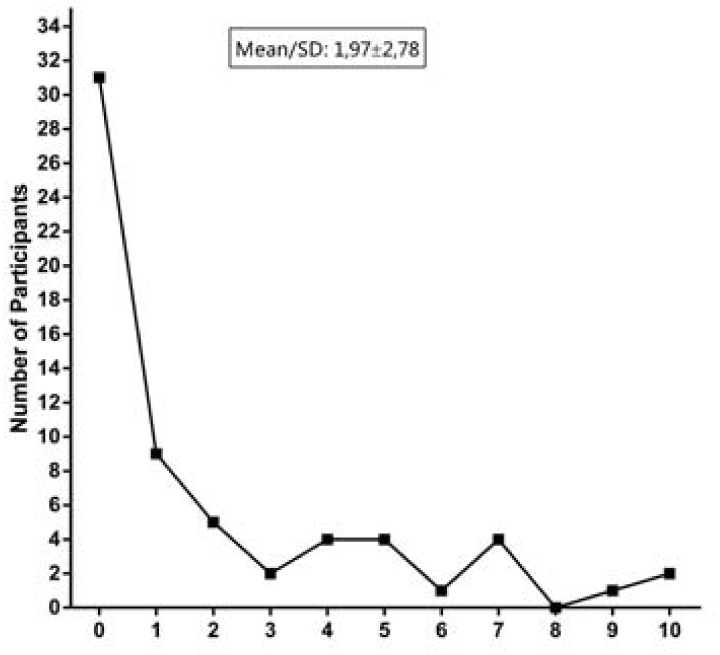
The presence of press members when performing CPR to a patient positively affects my CPR performance.

**Table 9.**
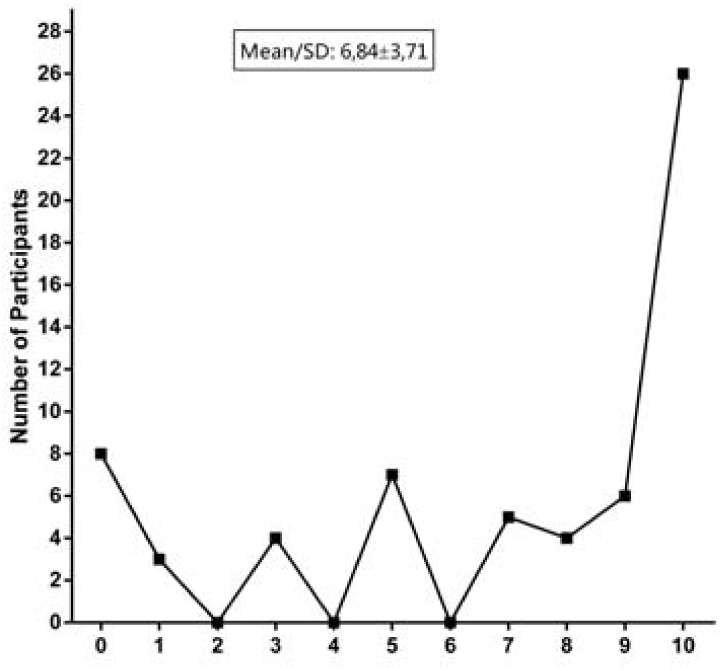
I think that taking image or shooting video by the press simultaneously when performing CPR on a patient affects the performance of the ambulance crew negatively.

**Table 10.**
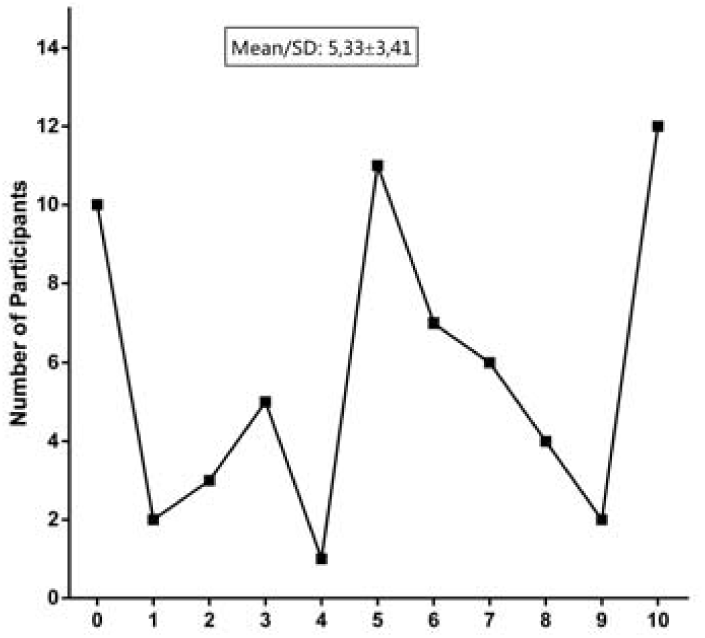
I understand some people’s desire to be with their loved ones at this difficult moment.

**Table 11.**
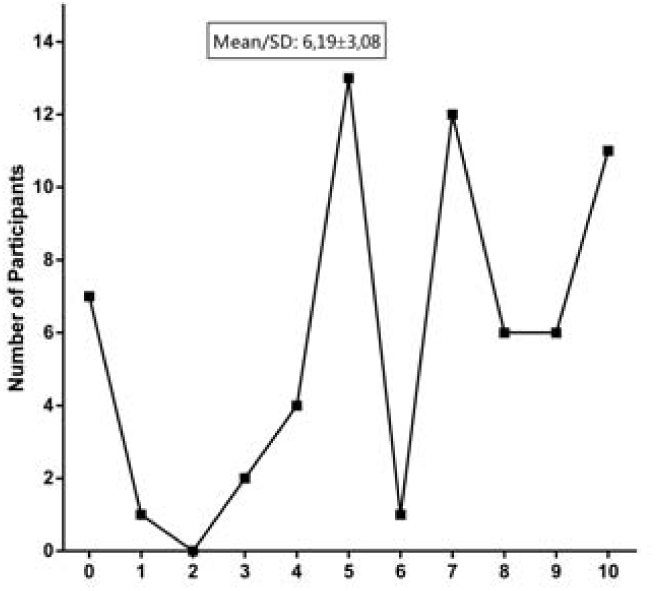
I think the main reason that a person wants to be there when CPR is applied to his relative is that he wants to see that everything should be done is really being done.

Significant differences in two phrases between gender is shown in Figure 1 and Figure 2. There is no significance between role and year of the experience.

**Figure 1.**
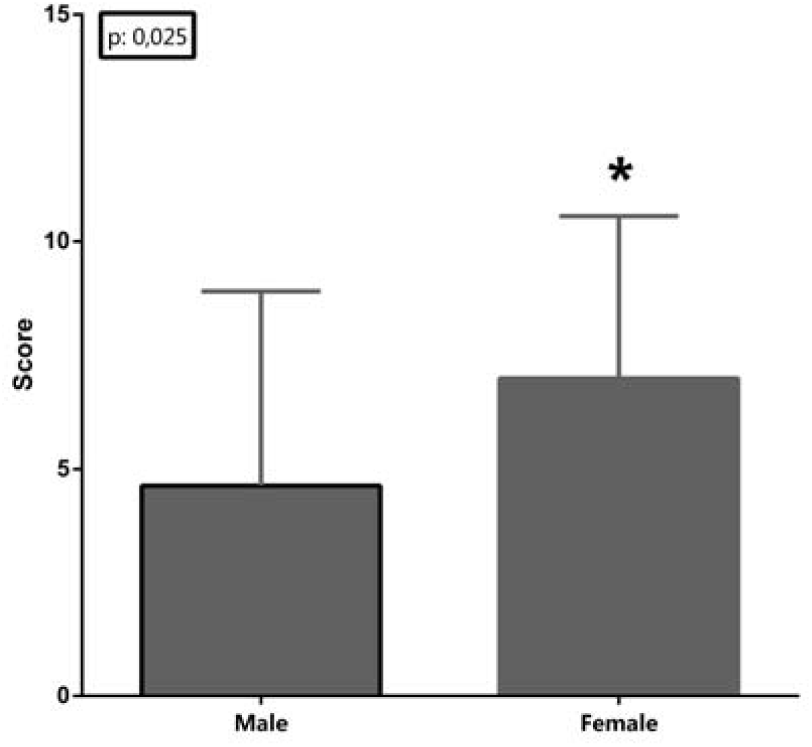
Significant difference in gender. (Mann Whitney U Test, p≤ 0,05) I think I would like to be there if I have the opportunity when being applied CPR to someone close to me.

**Figure 2.**
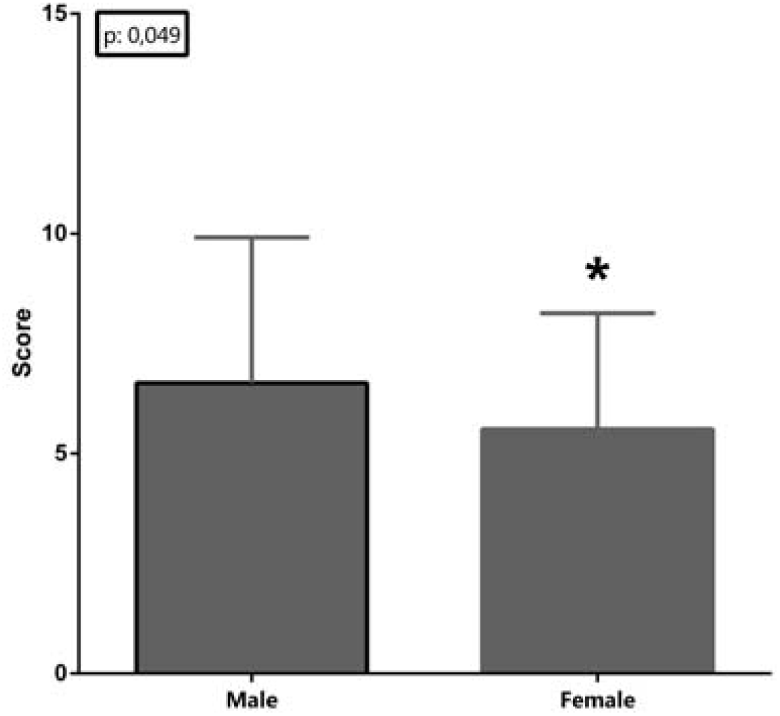
Significant difference in gender. (Mann Whitney U Test, p≤ 0,05) I think the main reason that a person wants to be there when CPR is applied to his relative is that he wants to see that everything should be done is really being done.

## Discussion

Pre-hospital emergency care is mostly different from hospital-based care. Because the proceeding here is generally acute and the decision-making process in this practice is generally both critical and has a complex nature[6]. CPR is one of the most combined processes in this field, as it includes medical, social, cultural and ethical issues. It is fast-paced, physical, intense, and often emotional for both health care professionals and bystanders[30]. This study focuses on the approach of pre-hospital emergency health care workers rather than the relatives. However, there is great variability in FPDR implementation in clinical practice.

The issue of allowing the presence of patients’ families by the health care providers is a paradigm shift[27,31]. Some health care professionals have positive attitudes toward FPDR[26,32–36], although some have concerns[37–40]. It is a pendulum oscillating between “benefits” and “disadvantages and/or limitations”[27]. The reasons for those who find FPDR favorable: Firstly, it is a “patient right” in terms of basic ethical principles[41–44], related to human dignity[5,45] and principle of “beneficence”[13,46,47]. Secondly, the simplicity to tell the relatives “what is going on”[12,17,33,48]. Thirdly, it makes easier for the relatives to accept the death and facilitates the grief process[17,49–51]. Fourthly, it provides the relatives feel well with loved ones in the last moments[33,34,52]. At the same time, there are many examples of both cultural and religion issues that independently support or oppose to FPDR[12,53–57]. On the other hand, the reasons for those who are against FPDR (in term of the negative attitude towards FPDR): Firstly, it increases stress burden in health professionals[58] and it may impact negatively on resuscitation attempt[36,59]. Secondly, it may include the risks of psychological trauma on relatives[5,22,24,56,60]. Thirdly, it may influence the decision-making process (i.e. the decision on termination of CPR or futile CPR)[7,61,62]. Fourthly, the risk of “communication problems” due to emotional stress of relatives[5,7]. The last reason is legal reservations[19,60,63].

In this study, PECs tend negative attitude to the FPDR. Of the respondents, 65.07% have strongly opposed to the family presence during CPR. Many other reasons for this negative aspect may be listed but, the most important reason for this finding could be that the justification of “being watched” which adversely affects CPR performance of the team. The thought of having relatives watching what PECs are doing at that time, is reasonably the main factor. For professional-based applications, such as CPR, it is important for PECs to be free from outside factors affecting the team performance. It is seen by the study findings that health professionals consider feeling of “being watched” to be a negative external effect for them. However, according to some studies PECs think they do not experience more stress when relatives are present because relatives do not interfere with the process[10,29,56]. This approach is probably due to two reasons; firstly the idea in PECs’ mind that the patient’s relatives do not intervene in the process, and secondly it is just experience. Therefore, the difference between two perceptions may be due to cultural and legal aspects which are changing country by country.

From the perspective of ethical conflicts experienced by PECs, it can be said that the conflict occurs between three broad principles; beneficence, nonmaleficence, and autonomy on FPDR. Giving family the autonomy to choose their involvement in such a critical moment is sure meaningful; however, there is an ethical conflict arises which is important; families’ autonomy and patients’ beneficence or maleficence? Although the basic principles of medical ethics appear to be accepted across cultures, the priority of these principles may vary among different cultures and ethics[64]. The principle here that PECs give priority is beneficence. The study states that at the point “being watched”, PECs’ CPR performance is adversely affected. Hence, it means that FPDR is not something beneficence for the patient. So, FPDR is not only the subject of medicine, but also sociology, culture and of course religion. This is probably the reason why many countries have different findings on the issue. Of course the perspective on end-of-life, death, and death-related issues are greatly influenced by cultural elements[65]. In that respect, the results of current study are the same with some studies results on the negative attitude of healthcare workers towards FPDR in Turkey[19,39,66,67].

This study reveals that PECs are not only familiar with the negative effect of the concept of FPDR on their team performance in particular, but also the negative effects of other PECs’ CPR performance. CPR performance is essential for linking patient to the chain of survival for treating cardiac arrest. However, the performance is highly variable both in out-of-hospital and in-hospital CPR[68,69]. The performance concerns in PECs’ mind may also create extra stress. Eventually, this negative approach may be due to increased stress burden in PECs during CPR and the atmosphere of the event. This anxiety also applies to possible violence and abuse[26]. PECs remain anxious over possible threats of violence and abuse from distressed significant others[7].

PECs generally perform CPR at the patient’s home and the effective communication with patient’s relatives is important during this process[40]. FPDR, especially in elderly patients, can have positive contributions in terms of accepting the impending death of the patient, communication and making possible contribution to health professionals[30]. However, it is very difficult to achieve good verbal communication, if only one ambulance team arrived at the home[5]. Because there are too many team-work to do systematically, repeatedly, and successfully for CPR attempt in a short time, but often there is only one PECs team. This situation is a challenge to manage and is one of the unique structures of the pre-hospital settings that include many conflicts[4]. Besides, systematic reviews report lower survival rates after cardiac resuscitation[70,71]. Thus, in a process that mortality is so high, immediate action is needed and time is limited, the approach of PECs to be on their own in the event is understandable. PECs always face to make a decision and they surely need to focus on what they are doing. In such a critical situation, effective communication may obviously not be among the priorities of PECs.

During a medical process that concerns one’s life, it is an ethical necessity to minimize all the elements that may have negative impacts on the performance of the professionals. It means that discussions on the FPDR need to be addressed in this respect. In this study, it can be thought that as the main reason for being there when applying CPR to someone close them; in the case they consider the possibility to help the CPR team there, as if they are the health professionals. At that point, they tend to see themselves not as ordinary relatives but as health professionals who know the technical details of CPR.

One of the prominent findings of this study is that PECs are strongly opposed especially to the presence of press members during CPR. Among the total sample, 71.41% of participants do not agree that presence of press members positively affects their performance. This finding can be related to the sense of “being watched”, too. Some CPR implementations in pre-hospital emergency medicine occur in public where the press members are likely to take images. CPR performed in such areas may also be referred to as Out-of-Home CPR (OHCPR). Of course, taking control over the environment for

CPR is very different from the hospital environment in OHCPR. At this point, the efforts of the press members to take images adversely affect the CPR performances of PECs. It is the highest rate of the study that PECs do not want any press around them when performing CPR on a patient. It may be due to both anxiety of “being watched” and legal concerns.

In some studies, it is stated that there are differences between attitudes of health professionals towards FPDR among the various groups of healthcare providers.[63] FPDR is voiced more often by nurses than the physicians[24,33,72–74]. Surprisingly, the current study does not reveal such a difference. The reason why there is no such difference among pre-hospital caregivers may be the healthcare in this setting provided as a team. Participating medical intervention in pre-hospital emergency medicine is not an individual practice but it is performed as a team. The specific dynamics of the pre-hospital emergency health may be the reason why there is no significant difference between the subgroups.

Ambulance personnel in Turkey should hold one of three professional: physicians, paramedics, basic emergency medical technicians with vocational education, and drivers mostly focus on pre-hospital care. The ambulance team, consisting of these three caregivers, intervenes together in all emergency cases. The literature has shown that medical staff, especially if they are junior, have negative attitudes toward FPDR compared to other health professionals[61]. In other words, it has been stated in some studies that as the age of the profession increases, the approach to FPDR is getting more positive[9,26,38,43,61,67,72,75]. The positive perception may be related to their experience on this field. However, there is no significance in this study between clinically experienced participants and less experienced colleagues. In participants, the periods of the experience are mostly 1-3 years. Therefore, the results are seen to be consistent with the literature. Education is an important key for the attitude change. When CPR providers are presented with FPDR education, their opinion-based beliefs may be modified to lower their guard against the issue, and to improve overall support of FPDR[6,76]. Therefore, education and professional experience can be defined as factors that could change the PECs’ attitudes toward FPDR in this regard.

According to the findings, there seems to be uncertainty in participants respond about two issues; First one, some relatives’ desire to be with loved one at the last moment. It is an indication about FPDR that at the very least it seems humane. Undoubtedly, PECs are also human beings and they seem to respond to this expression in ambivalence between their professionality and human aspects. They may want to use their professional knowledge and skills for their loved ones. They may have been indecisive because they contradict other expressions contained in the study. Second issue is to include social and legal concerns. Allowing family to see that everything possible has been done for their loved one is controversial for PECs. The prejudice towards patient relatives is a condition that prevents effective communication. PECs seem indecisive about what attitude they will take against people looking for deficiencies and inaccuracies in their transactions. CPR, when not properly initiated could have negative consequences on both PECs and significant others[43].

However, the study found significant differences in two phrases according to the gender. Women seem to be a more favorable presence with their loved one’s CPR than men. It may be one perception of women can meet the needs of PECs in that critical time by participating in the CPR attempt. This may also be the result of different coping skills between men and women in highly emotional processes. The second phrase includes a difference between genders is the intention of relatives to learn if everything is being done to their loved ones. Women are less favorable to this approach. As an implicit intention, it may be considered that relatives of patients have a desire to take control or observe the CPR process. Somehow, women seem to be less likely to accept this idea. Perhaps, considering these issues in terms of feminist bioethics may lead to more results that are descriptive.

In light of these results, PECs are opposed to the presence of both significant others and press during CPR. As they think their CPR performance is affected negatively, it is necessary to decrease such additional stress on healthcare workers. In today’s world, everyone is like a member of the press due to the widespread usage of social media. The probability of broadcasting in media of possible negative images creates stress for health workers on the same grounds. Consequently, this “audience pressure” makes a great effect on PECs during CPR. Additionally, “journalism concerns” should not interfere with the efforts of PECs “to provide qualified emergency healthcare”. Press organizations also have important duties in this regard. It is only possible to remove some of these obstacles in order to provide effective emergency healthcare with the ethical sensitivity of professionals in all areas. Therefore, it is necessary to provide an effective cooperation between health and press organizations and raise some sensitivity in this interaction.

Of course, FPDR can also be affected by the following structural situations which vary from country to country; the structure of the ambulance system in the country, the practice and legislation, the culture and cultural / individual differences, and the individual sensitivity and educational issues. Of all, in some cases in pre-hospital settings, family presence could facilitate better understanding among relatives. In the context of current study, PECs’ perceptions may be influenced by the lack of effective FPDR policy and guidelines in Turkey. Some additional studies need to be done and further works using larger samples in pre-hospital healthcare workers could find out more information on FPDR.

## Conclusion

On the basis of these findings, the presence of significant others during CPR affects pre-hospital emergency caregivers negatively. The disturbing effect on caregivers is related to not only the presence of family members or significant others but also the press and audience. Consequently, it includes performance anxiety among team members. Pre-hospital emergency settings are unique and this study shows the importance of ethics and sensitivity among pre-hospital caregivers who face to make ethical decisions in critical situations.

Family presence and audience effect are crucial issues that need more attention in pre-hospital cardiopulmonary resuscitation. Further research in this setting is necessary to explore the impact of family presence during cardiopulmonary resuscitation on emergency healthcare workers. It is suggested that more policy, support and educational intervention including ethics is required to provide pre-hospital caregivers with improved guidance of family presence during resuscitation. It is also necessary to provide effective cooperation between organizations of health and press to raise some sensitivity in this interaction.

## Data Availability

All data referred to in the manuscript is available. Data are available upon reasonable request.

## Study limitations

The study has several limitations. First, the study findings are based on a small sample of PECs; however, focused on an exact theme and adequate number of the phrases is likely to be sufficient to present the perceptions. Second, it is unable to measure the attitudes of healthcare workers directly on a controversial issue relating to ethics, law, religion, and culture. Third, collecting data through a questionnaire may have led participants to be unable to express themselves efficiently. Finally, since there are few studies in this unique setting, the questionnaire used in this study may have parts to be improved.

